# Approbation of Self-directed Learning by first year medical students: a mixed method study

**DOI:** 10.1101/2021.08.29.21261700

**Authors:** Bharti Bhandari, Deepti Chopra, Prerna Agarwal, Aprajita Panwar, Daljit Kaur, Tanvir K Sidhu

**Author notes:** **Corresponding Author:** Bharti Bhandari, Associate Professor, Department of Physiology, Government Institute of Medical Sciences, Greater Noida., E-mail –.

## Abstract

**Background:** One of the primary roles played by Indian medical graduates is to be a lifelong learner. For being a lifelong learner, the students should inculcate the habit of Self-Directed Learning (SDL). Lack of SDL skills among undergraduate medical students is a concern, hence the study was planned to introduce SDL in Physiology to phase-1 undergraduate medical students and assess its effectiveness through students’ and faculty’s perceptions.

**Methodology:** The project was commenced after obtaining clearance from the Institutional Ethics Committee. The faculty members and students were sensitized on SDL. Feedback questionnaire was framed and the topics for SDL were selected. Six topics were covered as part of the project. The effectiveness of the sessions was evaluated by administering the feedback questionnaire to the students and recording perceptions of the students and faculty on SDL. The quantitative and qualitative analysis of the data was done.

**Results:** A total of 96 phase-1 students participated in the study. Majority of the students felt that self-directed learning sessions have improved their SDL skills; they are more prepared and aware of their learning strengths and have started taking ownership of their learning. However some students felt the activity was not sufficient and SDL was not useful in improving their analytical skills. Both the students and the faculty were fairly satisfied by this teaching learning innovation.

**Conclusions:** Students and faculty were satisfied with the SDL strategy. SDL has shown to make them independent learner, who are aware of their learning goals and capable of evaluating their learning.

## Introduction

Fostering the attitude of ‘lifelong learning’ is now included as one of the roles of Indian medical graduate in the graduate medical regulations by National Medical Commission.^1^ ‘Lifelong learning’ means pliable and accessible learning that is carried out throughout the career of medical students.^2^ Untiring efforts have been put up by the medical teachers for transforming the medical students from passive to active learners, so as to create refined physicians who are capable of handling these evolutions in the medical education policies.^3^ The medical educators belief that the active learning strategies during undergraduate years may motivate the medical students in imbibing the habit of self-directed learning and help them in becoming lifelong learners.

Self-directed learning (SDL) is the latest educational approach embraced by the medical educators worldwide. SDL was described by Malcolm Knowles who believed that effective learning occurs in a self-directed and problem-oriented learning environment, and the student must realize a “need to know” and be genuinely self-motivated to learn.^4-6^The introduction of Competency Based Medical Education (CBME) by the National Medical Commission (NMC) has brought a paradigm shift in the medical education system in India. More emphasis is being given to student centered teaching-learning approach over teacher centered approach. The Self-directed learning sessions have become mandatory in each medical speciality in all the phases of medical undergraduate training. Self directed learning is a student centered approach, that would help the medical students to inculcate the habit of reading on their own and become lifelong learners. The latest Graduate Medical Regulations envisage being lifelong learner as one of the goals of Indian Medical Graduate. Lack of self-directed learning skills among undergraduate medical students is a concern. The current study was a part of FAIMER fellowship project, which was planned to introduce SDL sessions in Physiology to phase-1 undergraduate medical students. The short term objectives of the project were to implement SDL sessions to phase-1 undergraduate medical students in Physiology and to assess the acceptance of self directed learning as a learning strategy.

## Methodology

This quasi-experimental study was conducted in the Department of Physiology after obtaining approval from the Institutional ethics Committee. The participants were phase-1 medical students - 2019 **B**atch (n=100).

### Flow chart of methodology

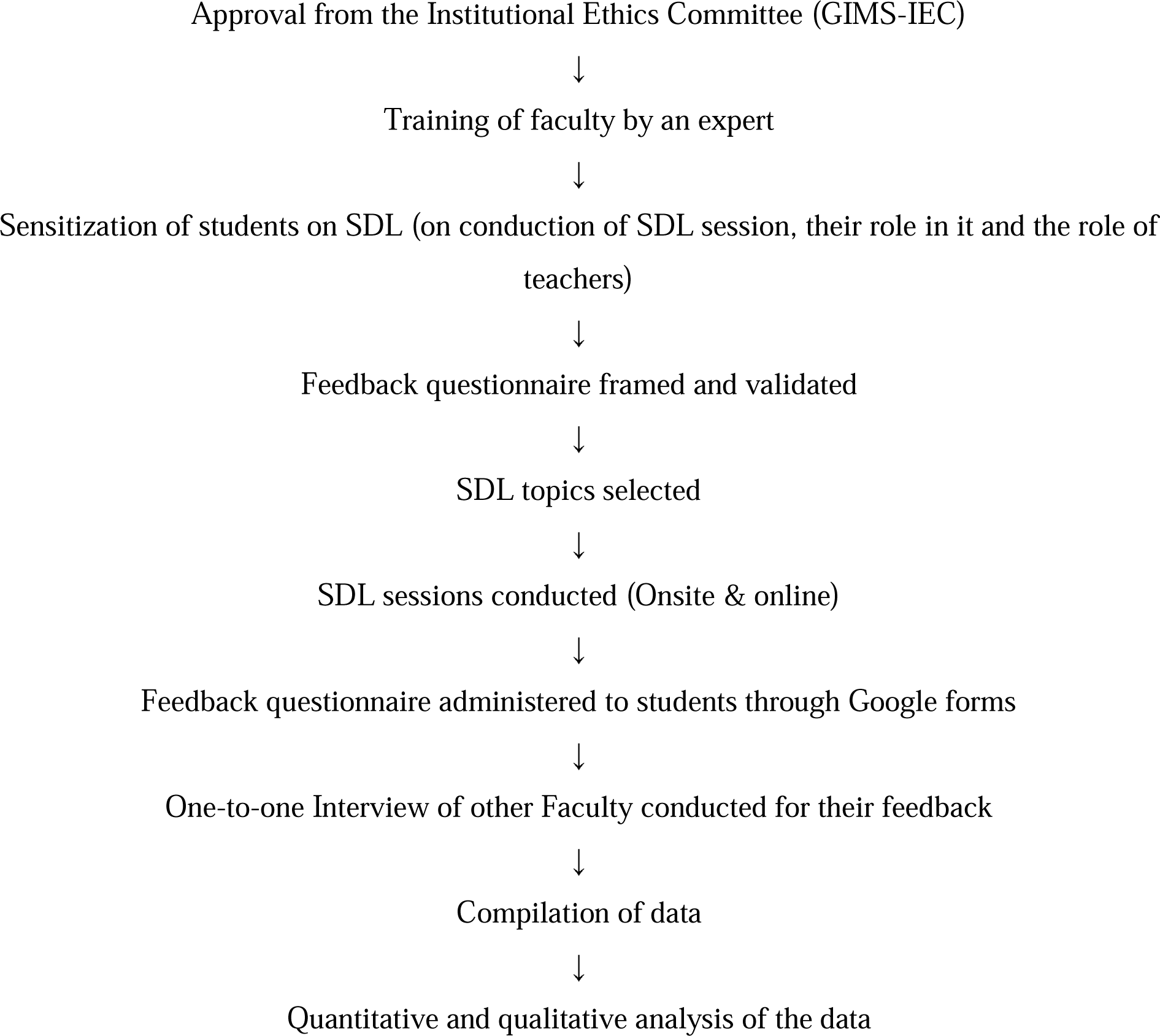

The Feedback survey questionnaire was prepared with the help of members of the Medical Education Unit of our institute. Open feedback was used to validate the questionnaire and it was revised accordingly. It was further modified and validated during the Progress Report session of FAIMER fellowship. The topics and schedule for SDL were finalized with inputs from the head of the department and other faculty. The topics were selected from the NMC curriculum for undergraduate students. These were – Blood banking and clinical importance of blood grouping; pathophysiology of myasthenia gravis; factors affecting heart rate; diffusion capacity of the lungs; hearing tests and primary hypothyroidism.

### Conduction of SDL sessions

Six SDL sessions were conducted, out of which, only four sessions could be conducted in the department, the remaining 2 sessions were conducted online. The students were informed in advance about the date and time of the SDL sessions. The topics were selected after through discussion with the fellow faculty. Each selected topic was covered in two sessions. Session 1 of 60 minutes duration included setting learning objectives, with the help of facilitators (if required) in small groups, finding appropriate learning resources with help from facilitators, if required and reading from books or E-resources (independently or collaborative). This self-study phase continued over the intervening period of 4 days. During Session 2 (120 mins duration), discussion was carried out in small groups as a way to self evaluate themselves. This was followed by reflection writing, in which they reflected on the learning process. Students were also assessed through MCQs test.

### Feedback

After completing six topics in SDL, feedback from the students was obtained. The questionnaire was shared online through Google form. Filling of the form was taken as implied consent. Before sharing the feedback form, the students were briefed about the importance of their honest and critical feedback. The feedback questionnaire had two types of questions: 1) questions with a 5-point Likert scale, and 2) open-ended question. In the 10 item questionnaire, the participants were asked to select from a Likert scale 5-point rating: “strongly disagree,” “disagree,” “neutral,” “agree,” and “strongly agree.”

For obtaining perceptions of the faculty, one-to-one interview was conducted using standardized open ended questions.^7^ The questions that were put up to the interviewee were - What are your views on the SDL sessions? How can it be improved further? What do you have to say about the students’ participation in the activity? What was your role during the conduction of the SDL sessions?

### Data analysis

#### Quantitative analysis (Descriptive statistics)

The responses of all the items on the Likert scale were tabulated and the satisfaction index for each item was calculated using the following formula^8^:

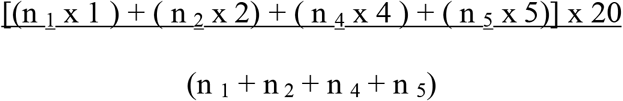

where, n is the total number of students gaining the score mentioned in the subscript for that particular item. Satisfaction index was calculated for each item to assess how satisfied the students were for each item, with the activity.

### Qualitative analysis

For recording perceptions of the students, open ended questions on Self-directed learning were asked using Google form. The students were asked about the usefulness of self-directed learning (insights), ways to improve it and factors facilitating and hindering the sessions. Faculty perceptions were obtained through one-to-one interviews. For the qualitative analysis, the responses to the open ended questions were reviewed, and thematically analyzed by two of the investigators (BB & TKS). Final agreement was reached after thorough discussions between them. The findings were clustered in groups: perceptions about SDL; suggestions for improvement, factors facilitating and hindering SDL.

## Results

A total of 99 students enrolled and participated in the self-directed learning activity for the project but feedback was obtained from 96 students only.

Feedback from students about the various aspects of the learning activity using a Likert scale is shown in Table 1. In the range from 1-100, the minimum satisfaction index of 65 was for item 6, which stated SDL improved the analytical ability; the maximum was 86 for items 2 and 7, which stated that the SDL engaged them in the learning process and had improved their ownership of learning. Figure 1 is the graphical representation of the satisfaction index.

**Table 1.**
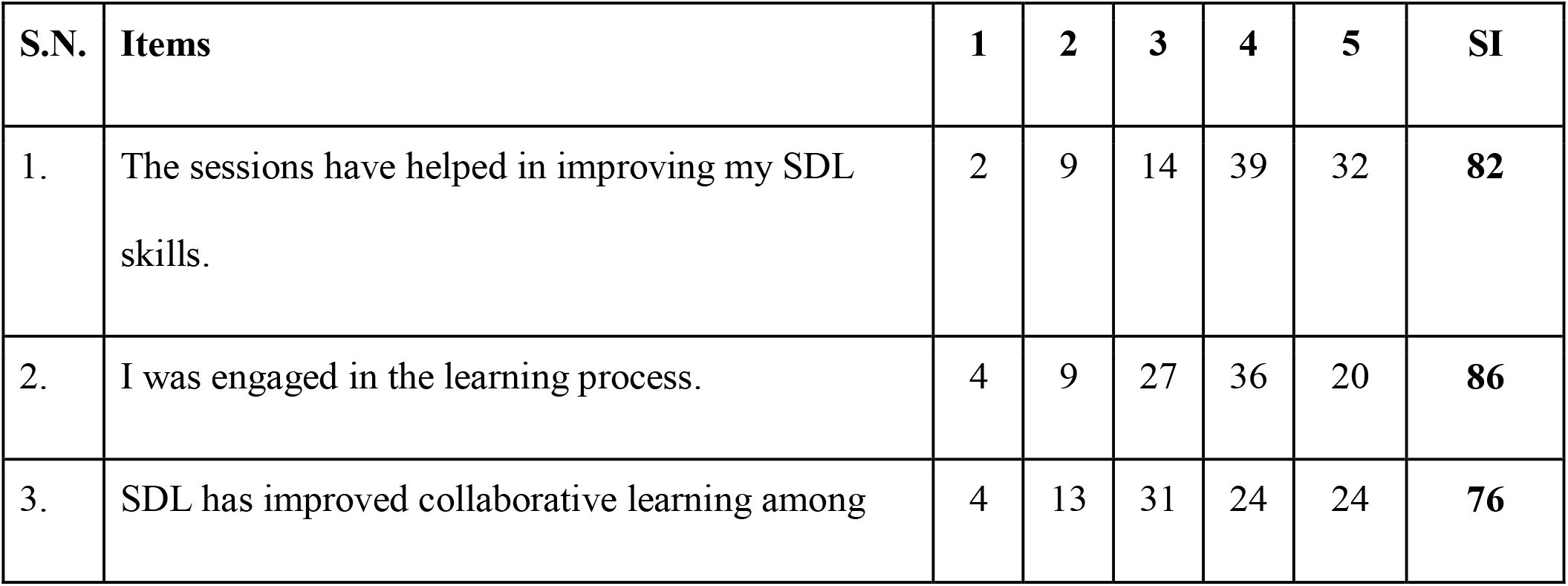
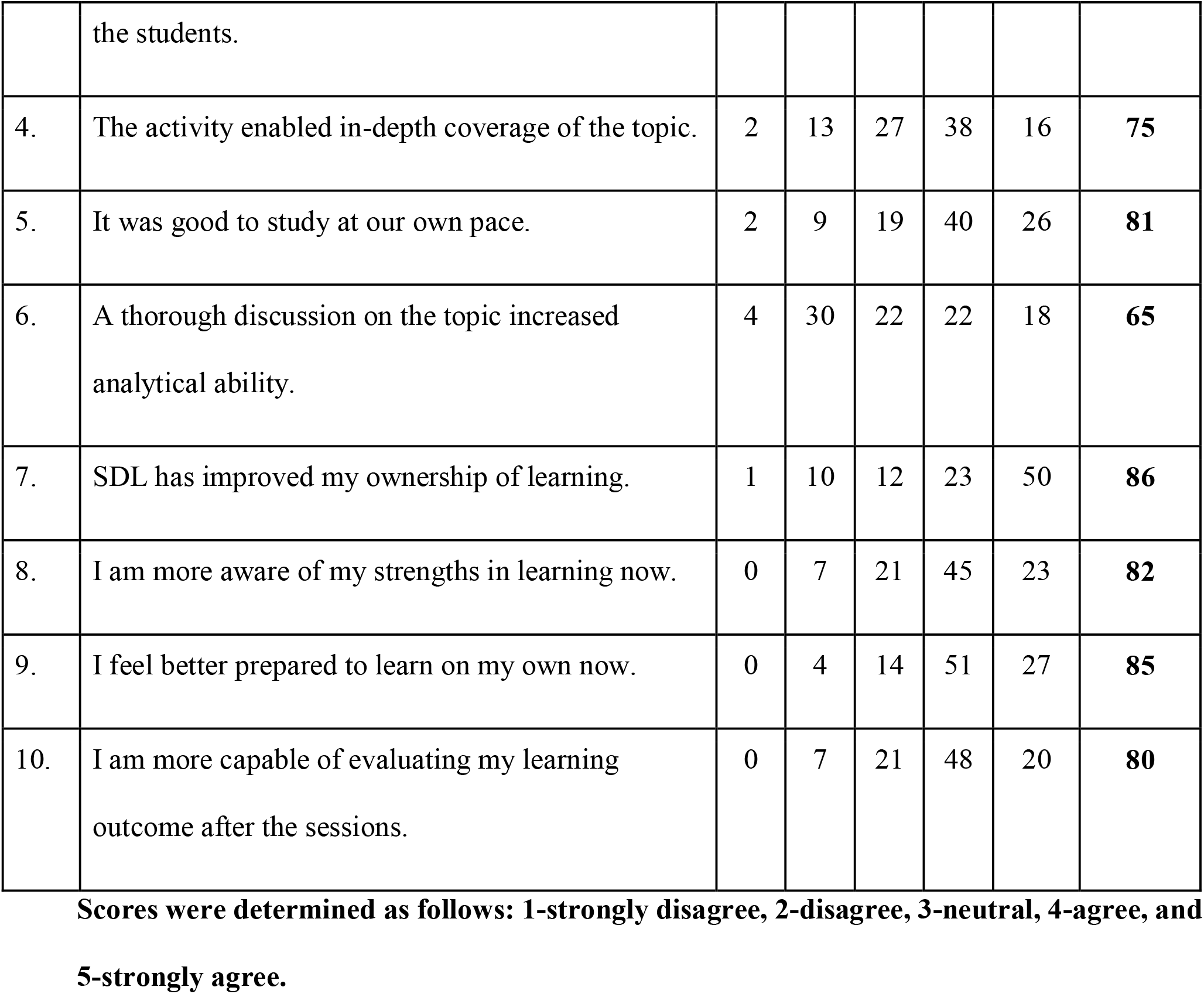
Student responses to the feedback questionnaire: scores.

| **S.N.** | **Items** | **1** | **2** | **3** | **4** | **5** | **SI** |
| --- | --- | --- | --- | --- | --- | --- | --- |
| 1. | The sessions have helped in improving my SDL skills. | 2 | 9 | 14 | 39 | 32 | **82** |
| 2. | I was engaged in the learning process. | 4 | 9 | 27 | 36 | 20 | **86** |
| 3. | SDL has improved collaborative learning among the students. | 4 | 13 | 31 | 24 | 24 | **76** |
| 4. | The activity enabled in-depth coverage of the topic. | 2 | 13 | 27 | 38 | 16 | **75** |
| 5. | It was good to study at our own pace. | 2 | 9 | 19 | 40 | 26 | **81** |
| 6. | A thorough discussion on the topic increased analytical ability. | 4 | 30 | 22 | 22 | 18 | **65** |
| 7. | SDL has improved my ownership of learning. | 1 | 10 | 12 | 23 | 50 | **86** |
| 8. | I am more aware of my strengths in learning now. | 0 | 7 | 21 | 45 | 23 | **82** |
| 9. | I feel better prepared to learn on my own now. | 0 | 4 | 14 | 51 | 27 | **85** |
| 10. | I am more capable of evaluating my learning outcome after the sessions. | 0 | 7 | 21 | 48 | 20 | **80** |

**Scores were determined as follows: 1-strongly disagree, 2-disagree, 3-neutral, 4-agree, and 5-strongly agree.**

Majority of students expressed that SDL was useful for better comprehension and it gave them a chance to explore newer ways of learning as per their styles. They could analyse their learning, improve their learning abilities and work on it further to improve the learning process. However, the students felt that such sessions should be planned targeting integration across the phases.

They expressed that SDL should be followed by students seminars or robust assessment on the selected topics to make it more effective. They expressed that a topic could be comprehended better through self-study but the role of the teacher cannot be ignored. Constant support and encouragement from the faculty members was of great help. Nonetheless, the interest of the students and the efforts put up by them made the SDL more effective. Among the hindering factors, the majority of students quoted poor network and internet connectivity and home environment as the primary factors during online sessions. They expressed that the sessions in quick succession also diverted their concentration.

The core ideas that emerged from open ended questions and some expressions are given in table 2.

**Table 2.**
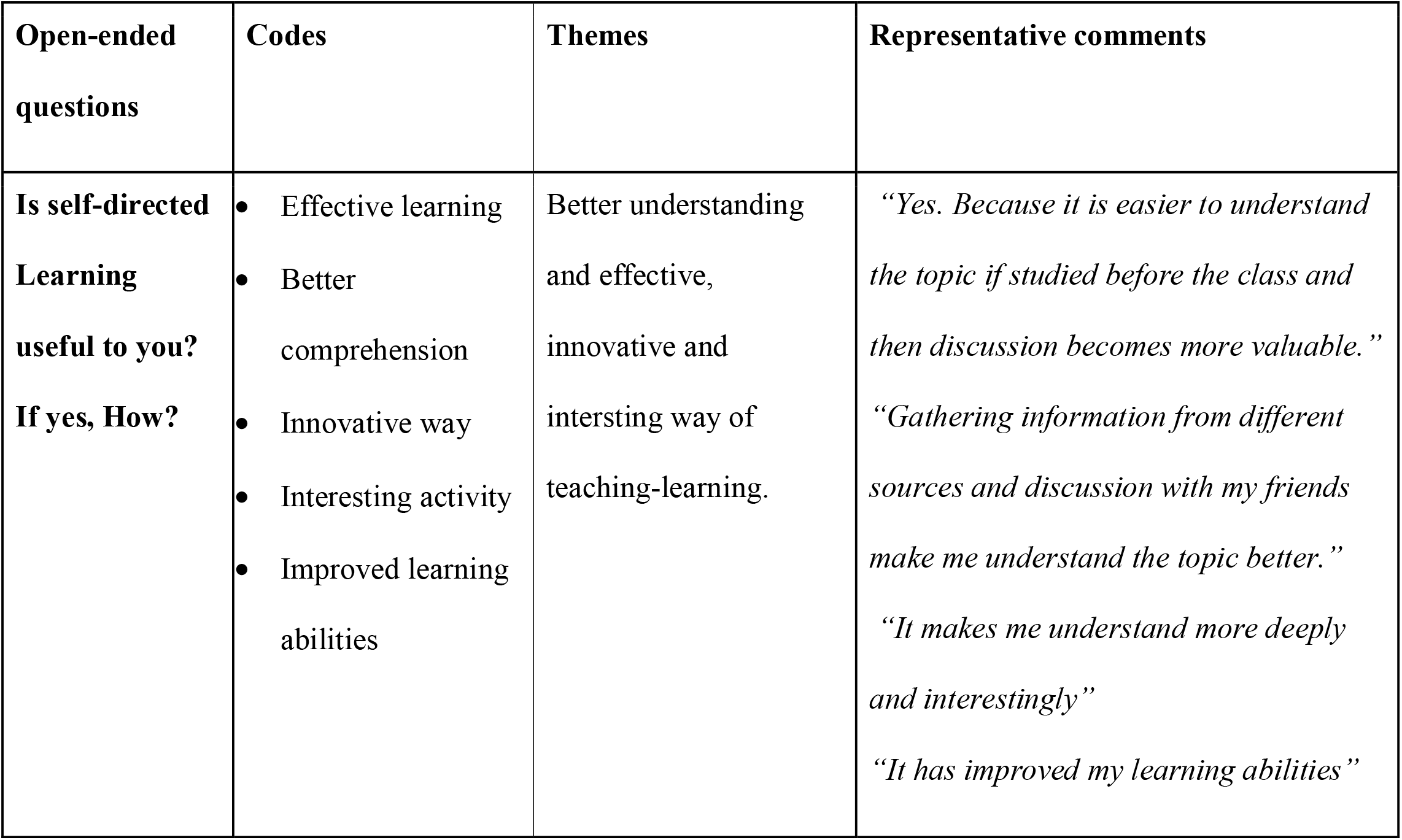
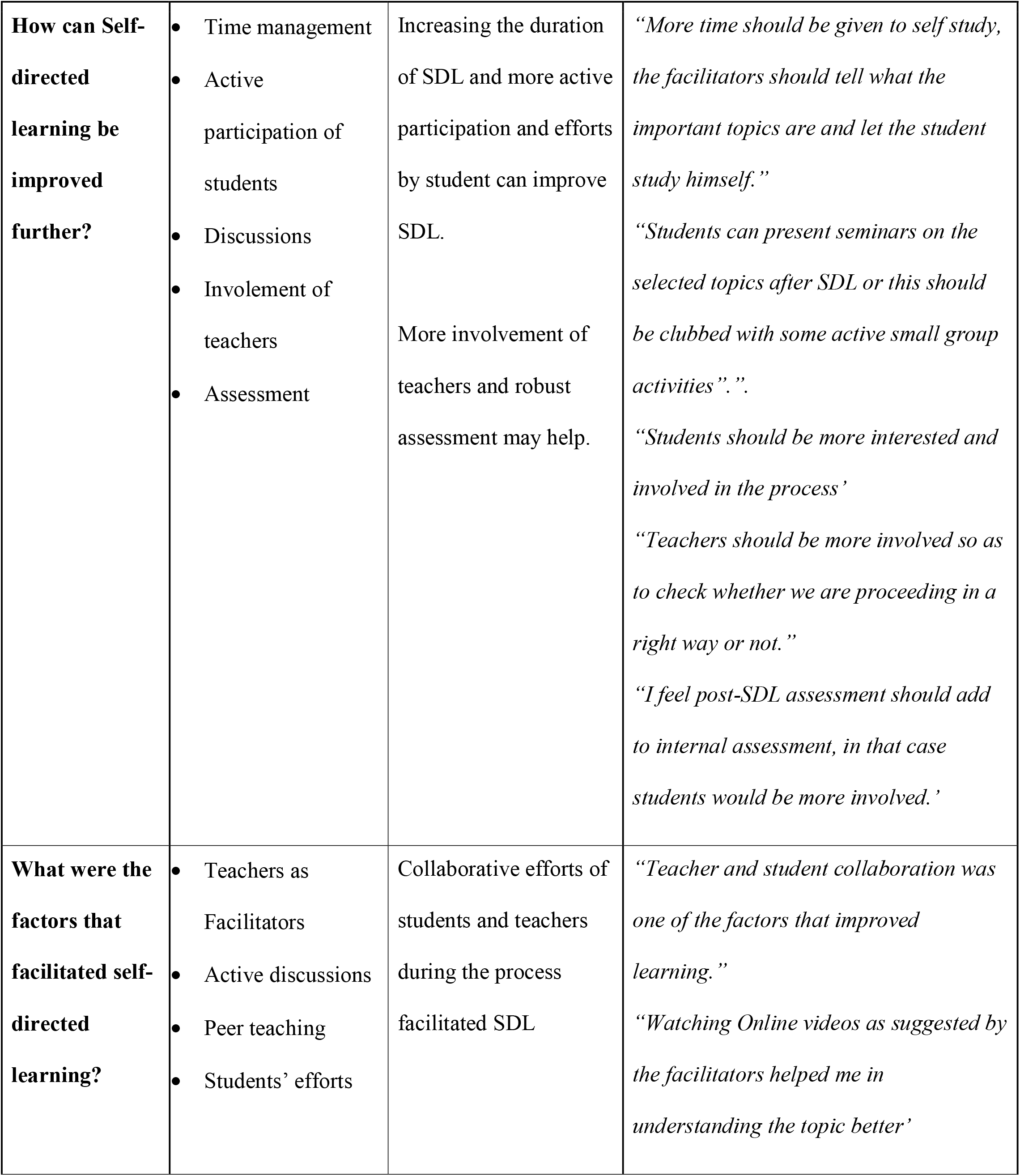
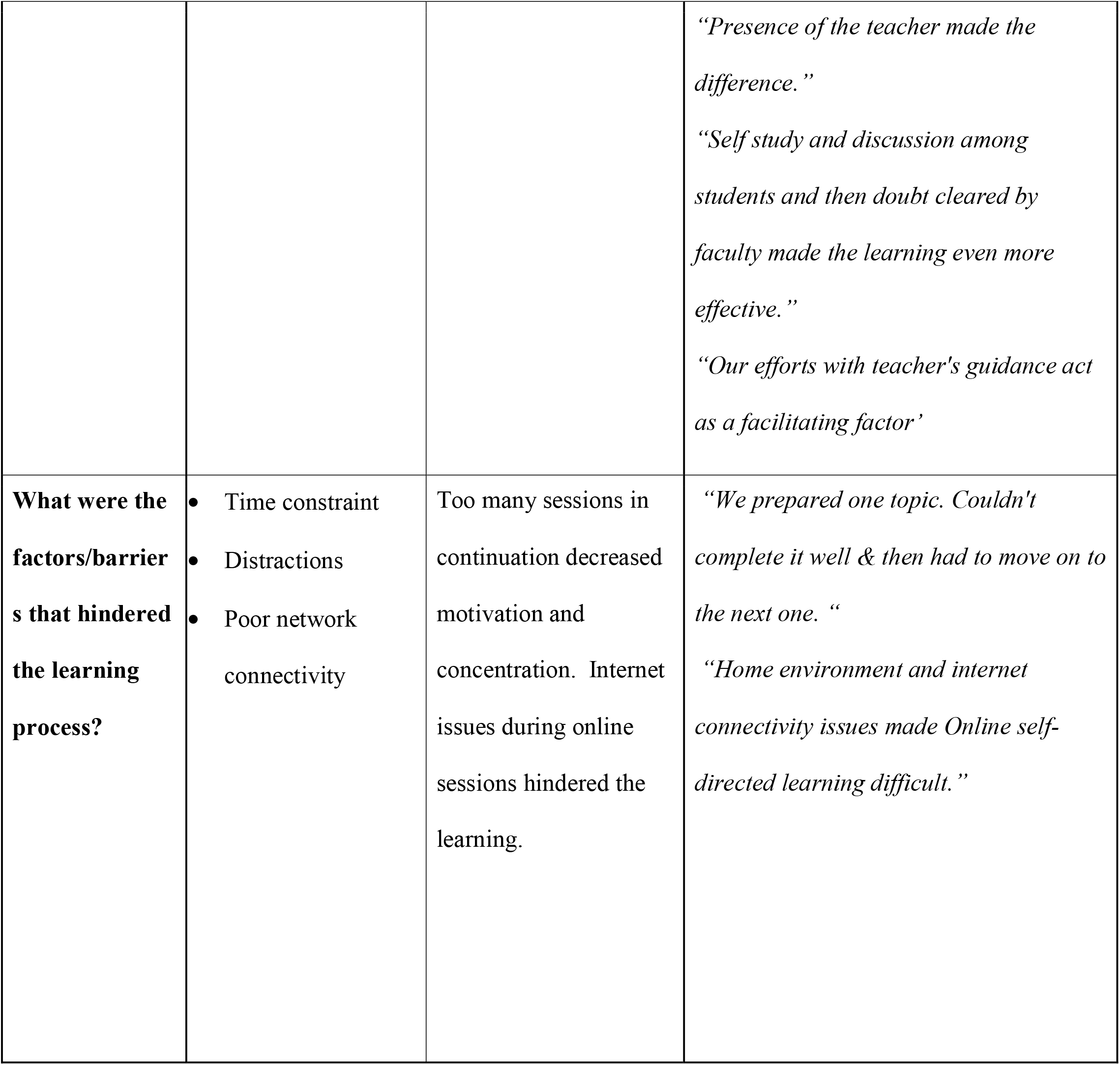
Themes that emerged in response to the open-question on the feedback questionnaire and representative comments from the students.

Only four faculty were involved in conducting the SDL sessions. They were interviewed using standardized open ended questions. The faculty views on SDL were very encouraging. They felt that SDL learning should be extended further to include some more topics but at the same time felt that conducting online sessions was a bit difficult as facilitators. The expressions from the interview on the same were “*I love conducting these sessions*.*” “I feel onsite sessions are more fun”*. As per the faculty, SDL can be improved by involving more number of facilitators. They thought that making groups as per the educational background and academic performance of the students would be better to ensure participation of all the students. “*Some students are very active and love to stay in the lime-light while some do not actively participate in the discussion*.*” “As per my views, grouping should be done taking into consideration their educational background and academic performances so that weak students can be motivated to come forward and participate*.*”*

All the faculty were satisfied with the way students had participated in the sessions. However there were always a bunch of students that never took initiative to be a part of the discussion. Facilitators expressed that giving topics before the sessions would be helpful for the weaker students. To ensure that the activity remains self-directed, the facilitators did not show their presence actively. They helped the students during formulation of learning objectives and suggesting learning resources, if required. They interfered only when the discussion was off track. Throughout the activity, they were silent observers. *“The most difficult part of the activity was to observe the students and not teach them :). “Ensuring participation of all the students was a difficult task “*.

## Discussion

The primary educational goal of this study was to provide opportunity to phase 1 medical students to develop skills essential for lifelong learning. SDL activities in small groups were planned for the medical students as per the guidelines of the National Medical Commission (erstwhile Medical Council of India).^1^ Various methods have been employed for developing self-directed learning skills among medical students by integrating the principles of SDL into team-based, case-based or problem-based learning.^9-11^ Similarly, such activities have also been planned using virtual patients.^12^

Our research question was to test the acceptability of SDL approach in phase-1 students and assess their perceptions on the same. The feedback from the students and faculty proved that the implementation of SDL activity in phase-1 students was achievable. Numerous studies have compared the acquisition of knowledge by students using the SDL approach with traditional methods of teaching.^13-15^ A systematic review by Murad et al had assessed the effectiveness of SDL in the health professions.^16^ They found that SDL activities moderately improved knowledge acquisition when compared to traditional teaching methods.

Acquiring SDL skills requires specific qualities in students like time management, stress management, being focussed and motivated, capability of identifying their learning goals and learning strategies to name a few. Presence of these skills helps students in acquiring self-directed learning skills.^17,18^ Although medical students possess most of these skills owing to their vast curriculum and the pace of the course, some of the students have identified lack of time and stress management skills and inter communication skills as some of the areas where they need improvement.^18^

In the current study, students scored high in most of the items of the feedback questionnaire, however they felt that SDL did not help them in improving their analytical skills. The students in our study were pre-clinical phase 1 medical students, they still need to develop the skills of integrating, reasoning and analyzing. This could be promoted through a case-based or problem-based SDL approach.^19, 20^ To improve this skill, most of the students felt that active participation of students is must and suggested integrating the activity across the phases. Discussion and reflective writing after self study phase helped students in identifying gaps in their knowledge. Reflection is critical in the training of future physicians as they develop clinical reasoning skills and provide evidence-based care.^21^ Asking students to reflect on the learning process during SDL was a valuable component of this activity because students were able to self-evaluate and identify their strengths and weaknesses.

Majority of students felt that the activity had instilled the quality of ownership of learning among them and it helped in better understanding of the topic. Students reported collaborating with peers during discussion of the topic. Forming small groups gave an opportunity for peer support. The advantage of team making in SDL has been very well explained by Hill et al, 2019.^22^

In our study, the satisfaction index for most of the index was more than 70. The students considered SDL as an effective way of learning (SI-85). In another recently published article, it was shown that 67% percent of students were satisfied and 66% reported as motivated to study the allotted topic further.^23^ A few students in this activity considered lectures better than SDL as they were unable to concentrate during self-study, similar to opinion of some students in another study was Patra et al, 2020.^23^

In response to the open-ended question on usefulness of SDL, the majority said that SDL is useful as it improves understanding and is an effective way of learning. SDL gives them avenues to discover their styles of learning. The statements *“It is easy to understand the topic if read by oneself and then followed by discussion”*; “*I can analyse my learning style and explore my strengths*” reflect their feelings.

Students expressed that SDL can be improved by robust assessment, by integration, by choosing the topics judiciously and by increasing its duration. They felt that active involvement of students in the form of seminars may also improve SDL skills. *“A list of important topics from the chapter should be given, which then should be prepared by the students*.*”“Horizontal and vertical integration will make more understandable” “It can be more analytical and practical*.*”* These are some excerpts that express their needs. It has been shown by researchers that appropriate use of SDL may increase future clinical practice.^24^

Majority of the students expressed that motivation by the facilitators was very encouraging. Additionally discussion with their peers helped a lot. However they accepted facilitators to participate more in suggesting reading resource materials. Contrary to this, Murad et al (2010), postulated that SDL was more effective when learners were involved in choosing learning resources.^16^ The reason for this contradictory perception of the students could be the level of our students, being phase-1 students, they needed more help during the activity. Another study also showed that the students face difficulty in finding the correct learning resources.^18^

Students wanted seminars and other such activities to be conducted as a part of the SDL session to promote active participation of all the students. It has been shown that a change in learning and behaviour in medical students is evident with conduction of such activities.^25^

Expecting students in the pre-clinical year to be entirely self-directed is too high an expectation. Developing self-directed learners is a gradual process, where teachers must facilitate the acquisition of skills required for SDL. As per Kidane et al, components of the hybrid curriculum, mainly PBL, could encourage preclinical students’ self-directed learning, as the curriculum is still not free from teacher-centred culture as the majority of teachers still have high power in deciding the learning process.^26^

The main hindrance quoted by most of the students was poor internet connectivity. Under such adverse circumstances, all sessions could not be conducted onsite. However as suggested by other researchers, internet-based learning is associated with large positive effects compared with no intervention.^27^

The strength of our study is the high response rate to open-ended questions. The weakness is that it was conducted in one institution only. The Feedback obtained through questionnaire is subjected to response bias, primarily acquiescence bias.

The medical teachers should not believe that medical students have all the skills required for engaging them in SDL activities. We recommend evaluation of the readiness of incoming medical students for SDL activities and evaluation of their SDL learning abilities. The students should be sensitized to the process of self directed learning and there should be gradual implementation of activities that enhance their SDL skills.

## Conclusions

The study has shown that SDL is well accepted by the majority of the students and faculty and they were satisfied with the approach. It has shown to improve the understanding of the subject and the students feel better prepared for learning on their own. This study also demonstrates that formal SDL sessions can be successfully implemented in Physiology and may contribute to the lifelong learning skills that are pertinent for improving patient care.

## Data Availability

Data would be made available by the corresponding author on request.

